# Development and Validation of a web-based Postoperative *Clostridioides difficile* infection risk prediction model

**DOI:** 10.1101/2020.06.23.20138420

**Authors:** Sang H. Woo, Bryan Hess, Lily Ackermann, Scott W. Cowan, Jennifer Valentine

**Affiliations:** Department of Medicine- Division of Hospital Medicine, Thomas Jefferson University, Philadelphia, PA 19107; Department of Medicine- Division of Infectious Disease, Thomas Jefferson University, Philadelphia, PA 19107; Department of Surgery, Thomas Jefferson University, Philadelphia, PA 19107

## Abstract

**Background:** *Clostridioides difficile* infection is associated with significant morbidity, mortality and increased costs. Assessment of the postoperative *C. difficile* infection risk is necessary to improve the outcome of surgical patients.

**Objective:** To develop and validate a risk prediction tool for *C. difficile* infection after surgery.

**Methods:** In this retrospective cohort study, 2,451,169 surgical patients from the American College of Surgeons National Surgical Quality Improvement Program Database (ACS-NSQIP) over 2015-2017 were included. Nine predictors were selected for the model: age, preoperative leukocytosis (>12 ×10^9^/L), hematocrit (≤30%), chronic dialysis, insulin dependent diabetes, weight loss, steroid use, presence of preoperative sepsis, and surgery type. A second model included hospital length of stay as a predictor. A predictive model was developed using ACS-NSQIP 2015-2016 training cohort (n=1,435,157) and tested using 2017 validation cohort (n=1,016,012). Multivariate logistic regression was used for the model.

**Main outcome:** The primary outcome was postoperative 30-day *C. difficile* infection (CDI).

**Results:** 0.39% of the patients (n=9,675) developed CDI and 42.3% (n=4,091) of CDI occurred post-discharge. The *Clostridioides difficile* risk prediction model had excellent AUC (area under the receiver operating characteristic curve) for postoperative *C. difficile* infection (training cohort=0.804, test cohort= 0.803). The model that includes hospital length of stay has a high AUC (training cohort=0.841, test cohort=0.838).

**Conclusion:** The *C. difficile* prediction model provides a robust predictive tool for postoperative *C. difficile* infection.

## Background

*Clostridioides* (*formerly Clostridium*) *difficile* is a commonly identified cause of healthcare-associated infection in the United States.^1^ Preventive measures include improving antibiotic use, healthcare facility cleaning, patient isolation, and hand hygiene. ^2–4^

Despite aggressive preventive measures, the clinical and economic impacts of *C. difficile* infection (CDI) remain significant and *C. difficile* maintains an “Urgent Threat” status according to the Centers for Disease Control and Prevention (CDC). ^4–6^ There is a significant healthcare burden with *C*.*difficile* infection with increased morbidity and higher healthcare costs. ^3,6,7^ The CDC estimated 223,900 CDI cases in hospitalized patients in 2017, with an estimated 12,800 deaths and 1 billion dollars in estimated attributable healthcare costs. ^4^ In the same report, they noted that healthcare-associated CDI cases are decreasing, however community-associated cases are not. ^8^

The ability to predict and target specific *C*.*difficile* infection preventive measures to the highest risk patients has important implications for hospitals. In one large cohort of postoperative patients over a four-year period, CDIs developed at a rate of 0.4% per year, with a 12-fold increase in morbidity and 5-fold increase in mortality associated with CDI compared to post-op patients without CDI. ^9^ It was reported that over 8% of admitted patients are carriers of *C. difficile* with high risk of infection.^10^ Surgical patients are at particularly high risk for CDI. Extended antimicrobial prophylaxis given to reduce surgical site infection is associated with increased *C. difficile* infection. ^11,12^ There is also decreased throughput in hospitals and decreased availability of beds as CDIs necessitate a need for isolation and effective terminal cleaning processes. The Centers for Medicare and Medicaid Services uses hospital-acquired CDI rates as part of their reimbursement to hospitals and the financial penalties for institutions failing to meet quality metric standards are significant. ^13^ Even though *C. difficile* infection has been associated with significant morbidity and mortality ^6,14,15^, to our knowledge, a practical risk model for postoperative CDI based on a large number of patients and hospitals is not available. Oh et al. demonstrated a machine learning algorithm to predict the risk of CDI but it was trained on patients from only two hospitals. Additionally, its use of over 4000 variables limits ease of use in clinical settings and the model is not specific to surgical patients.^16^ Other studies showed predictive models for recurrent CDI with variable discrimination, but these were not for initial CDI infection prediction and not limited to surgical patients. ^17,18^

Our study was conducted using the database from the American College of Surgeon’s National Surgical Quality Improvement Program (ACS-NSQIP). ACS-NSQIP clinical data are collected from more than 500 US hospitals and includes preoperative clinical features, risk factors, and 30 day postoperative outcomes. ^19–21^

## Methods

### Study population

Data from 2015 to 2017 ACS-NSQIP database were used for the analysis. 2,451,169 participants were included in the study cohort. Study participants flow chart is shown in **eFigure I** in the **Supplement**. Clinical data and occurrences of complications were collected by trained surgical clinical reviewers at each participating hospital and participants were followed for 30 days after surgery. Analyzed risk factors included age, sex, smoking, history of congestive heart failure, history of chronic obstructive pulmonary disease, diabetes, presence of preoperative sepsis, use of hypertension medications, steroid use, admission origin, ascites, chronic dialysis, bleeding disorder, disseminated cancer, dyspnea, emergency surgery, preoperative renal failure, weight loss, American Society of Anesthesiologists’ class, and preoperative functional status. Preoperative laboratory tests in the analysis include serum sodium, creatinine, white blood cell count, and hematocrit.

### Outcome

The primary outcome was the occurrence of 30-day postoperative *Clostridioides difficile* infection. In the NSQIP, the diagnosis of CDI is met by one of two criteria: “documentation in the medical record of a positive *C. difficile* test result in the 30 day postoperative period, or documentation that the patient is receiving current treatment for *C. difficile*”. ^22,23^

### Data Analysis

2,451,169 patients from 2015-2017 were included in the initial analysis. Patient demographic factors were assessed using Pearson χ2 for categorical variables and Wilcoxon for continuous variables.

Our *C. difficile* predictive model was trained using data from years 2015 and 2016 (N= 1,435,157) and the model was tested using data from 2017 (N=1,016,012). Model predictors were chosen based on a backward elimination method and clinical importance. Multivariate logistic regression analysis was done to obtain predictor coefficient, standard error, and adjusted Odds ratio.

GridSearchCV package with 5-fold cross validation was used to select the model parameters. We used AUC (Area Under the Curve) to evaluate the performance of a model. Separate categories were made for missing values of laboratory concentrations (serum hematocrit, white blood cell count and creatinine). Flow diagram of the study cohort is in the online Supplement Figure I. Python (version 3.6.6), Statsmodels (version 0.9.0), r programming (RStudio version 1.1.463) were used for statistical analysis. Scikit-learn package was utilized for machine learning modeling and a web-based risk model. The Thomas Jefferson University institutional review board approved this study and waived informed consent from study participants.

### Results

## Results

### Participant Characteristics

The study included 2,451,169 participants from 2015-2017 cohort and 56.8% were women. The overall 30-day post-operative *C. difficile* infection incidence was 0.39% (n=9,675).

**Table 1** shows demographic characteristics of study participant cohorts from 2015-16. CDI rate was 0.42% (n=6,095). Patients who developed *C. difficile* infection were older (64.6 vs 56.5, p<0.001) and had a higher rate of insulin dependent diabetes (12.0% vs 5.8%, p<0.001). More Patients with C. *difficile* infection had disseminated cancer, COPD, and ESRD(p<0.001).

**Table 1.** Characteristics of the patients (patients from 2015-16)

|  | No *C. difficile* Infection<br>(n=1,429,062) | *C. difficile* infection<br>(n=6,095) | p-value |
| --- | --- | --- | --- |
| Age (Mean±SD) | 56.5±16.8 | 64.6±15.7 | * |
| Sex (Male) | 43.36 | 45.96 | * |
| Preoperative sepsis- None(%) | 94.65 | 79.34 | * |
| Preoperative SIRS(%) | 2.93 | 8.42 | * |
| Preoperative Sepsis(%) | 2.05 | 9.14 | * |
| Preoperative Septic Shock(%) | 0.36 | 3.1 | * |
| No diabetes(%) | 84.42 | 78.87 | * |
| Non-Insulin dependent diabetes(%) | 9.82 | 9.12 | * |
| Diabetes - Insulin dependent(%) | 5.76 | 12.01 | * |
| Hypertensive medication use(%) | 44.78 | 59.44 | * |
| Ascites(%) | 0.31 | 1.76 | * |
| Weight Loss(%) | 1.18 | 5.35 | * |
| Emergency surgery(%) | 8.84 | 21.2 | * |
| COPD(%) | 4.39 | 9.94 | * |
| Congestive Heart Failure(%) | 0.87 | 3.43 | * |
| Disseminated cancer(%) | 2.26 | 6.43 | * |
| Smoker(%) | 17.64 | 20.34 | * |
| Preoperative renal failure(%) | 0.34 | 2.18 | * |
| Chronic dialysis (%) | 1.28 | 5.07 | * |
| Steroid use (%) | 3.59 | 8.11 | * |
| Bleeding disorder(%) | 4.04 | 11.88 | * |
| ASA Class 1-No Disturb(%) | 8.74 | 2.13 | * |
| 2-Mild Disturb(%) | 44.8 | 21.2 | * |
| 3-Severe Disturb(%) | 40.38 | 54.82 | * |
| 4-Life Threatening(%) | 5.9 | 20.75 | * |
| 5-Moribund(%) | 0.17 | 1.1 | * |
| Preoperative WBC ($\times 10^9/L$) | 8.15 | 9.76 | * |
| Preoperative Hematocrit(%) | 39.73 | 36.37 | * |
| Preoperative Sodium(mEq/L) | 139.08 | 138.25 | * |
| Preoperative Creatinine(mg/dL) | 1.00 | 1.30 | * |

\* P<0.001

### Postoperative 30-day mortality and readmission

**Figure 1** illustrates 30-day unadjusted mortality and readmission rates with postoperative *C. difficile* infection from 2015 to 2017. Patients who developed postoperative CDI had a high 30-day mortality rate (5.7% vs 0.9%). 30-day Readmission rates with CDI were also significantly higher (32.5% vs 5.1%).

**Figure 1.**
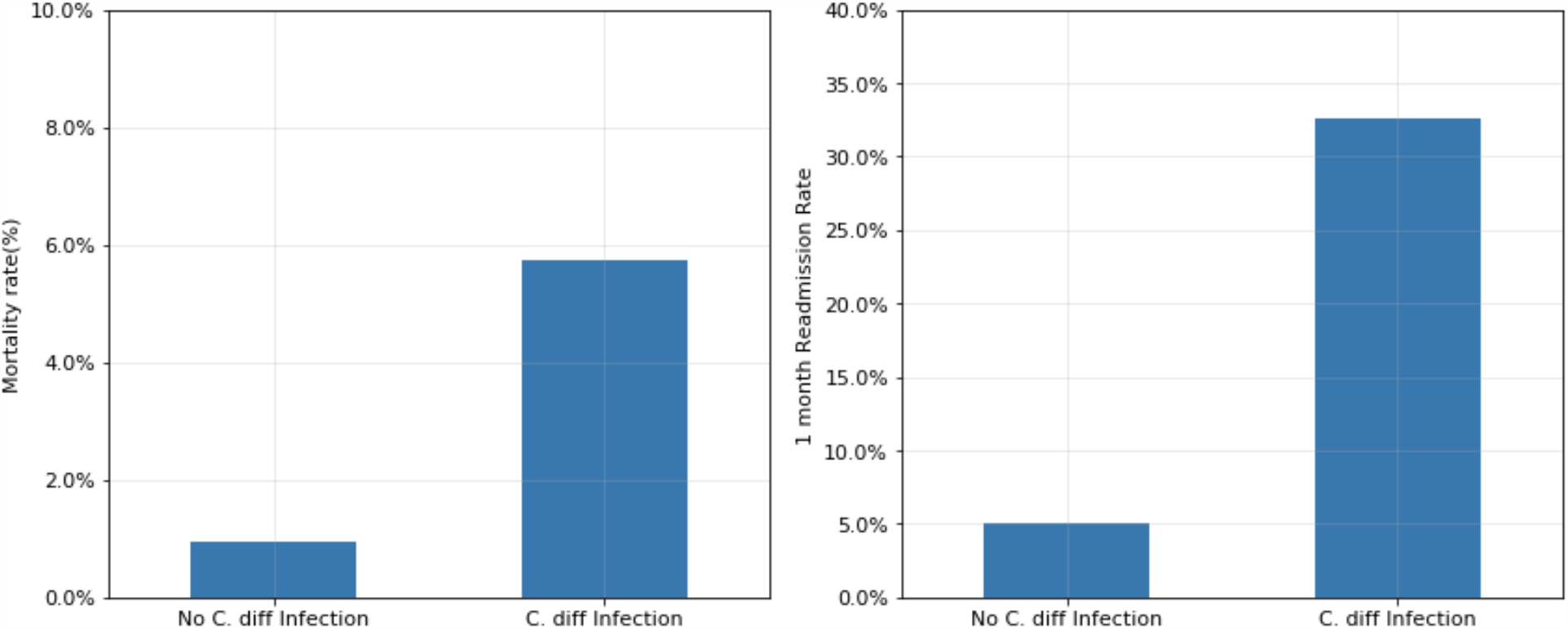
Unadjusted 30-day postoperative mortality and readmission rates associated with *C. difficile* infection.

### Age, leukocytosis, anemia, length of stay and incidence of *C. difficile* infection

**Figure 2** shows the unadjusted rates of postoperative *C. difficile* infection with age, preoperative leukocytosis (white blood cell count>12×109/L), preoperative anemia (hematocrit≤30%) and hospital length of stay from 2015-17 cohort. The incidence of infection increased with older age. Preoperative leukocytosis (0.9% vs 0.4%) and anemia (1.6% vs 0.4%) were significantly associated with CDI. Longer hospital length of stay also had significant association with the incidence of *C. difficile* infection (>10 days: 2.9%, 6-10 days:1.1%, 0-5 days 0.2%).

**Figure 2.**
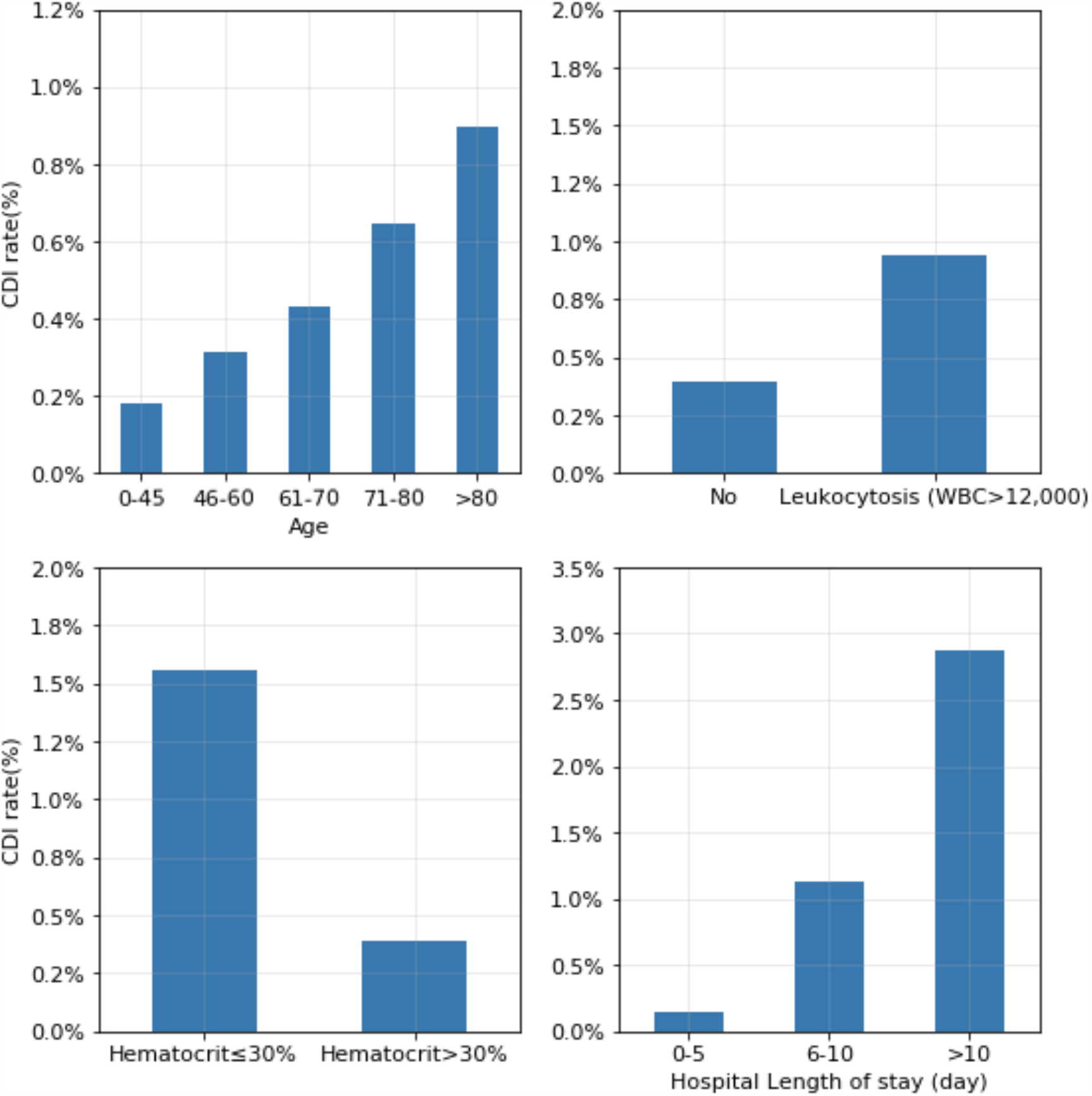
Age, Leukocytosis, Anemia and Hospital Length of Stay and *C. difficile* infection.

### Surgery type and *C. difficile* infection

There is a significant difference in the *C. difficile* infection rates between the types of surgery. **Figure 3** shows unadjusted CDI rate according to the type of surgery from the 2015-17 cohort. Intestinal surgery (1.56%), aorta vascular surgery (1.41%), and liver-pancreas surgery (1.34%) demonstrated particularly high rates of CDI. Thyroid surgery (0.05%) and orthopedic upper extremity (0.06%) demonstrated lower rates of CDI.

**Figure 3.**
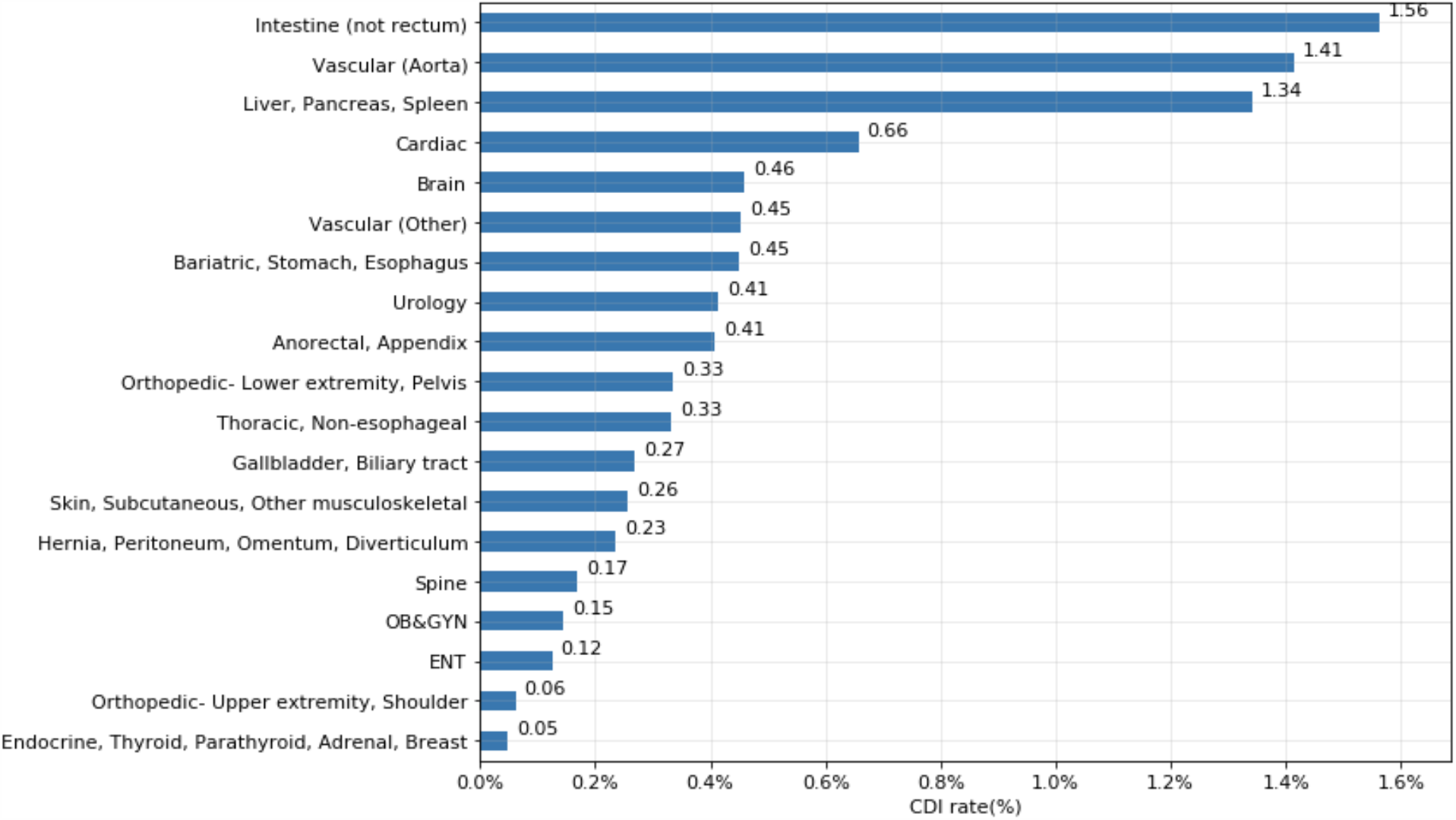
30-day postoperative *C. difficile* infection occurrences (%) according to surgery type.

### Prediction and validation of 30 day postoperative *C. difficile* infection

**Table 2** shows nine predictors of postoperative *C. difficile* infection. Age, preoperative leukocytosis (white blood cell count>12×109/L), preoperative anemia (hematocrit≤30%), chronic dialysis, preoperative sepsis, chronic corticosteroid use, and weight loss (>10% body weight loss in last 6 months) were significantly associated with CDI. Insulin dependent diabetes mellitus was also significant similar to a previous study. ^24^ Preoperative SIRS (Odds ratio (OR)=1.89, p<0.001), sepsis (OR=2.16, p<0.001), and septic shock (OR=2.25, p<0.001) were strongly associated with *C. difficile* infection. The model built with the training cohort was validated on the testing cohort of 1,016,012 patients from 2017. **eFigure II** in the **Supplement** shows excellent calibration, well matched with the 45-degree line.

**Table 2.**
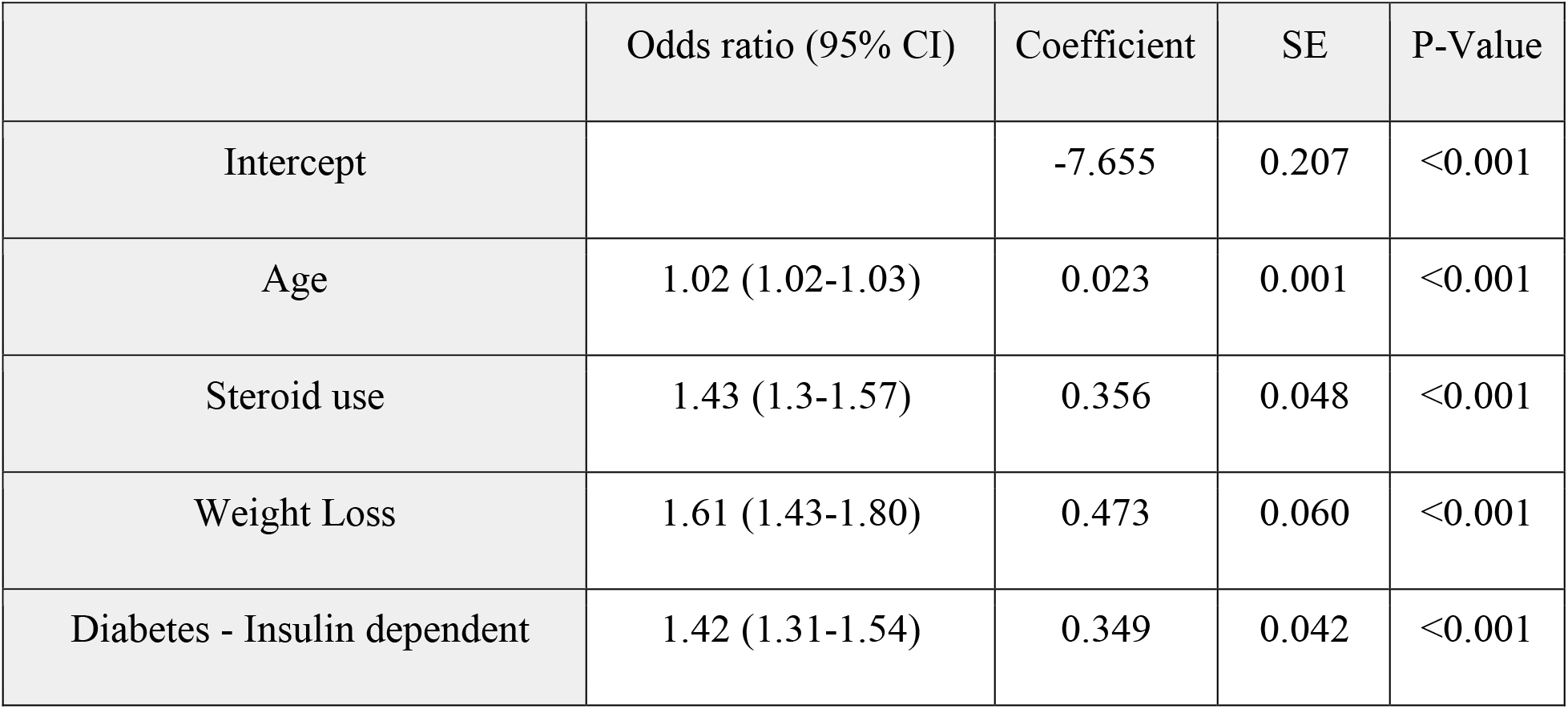
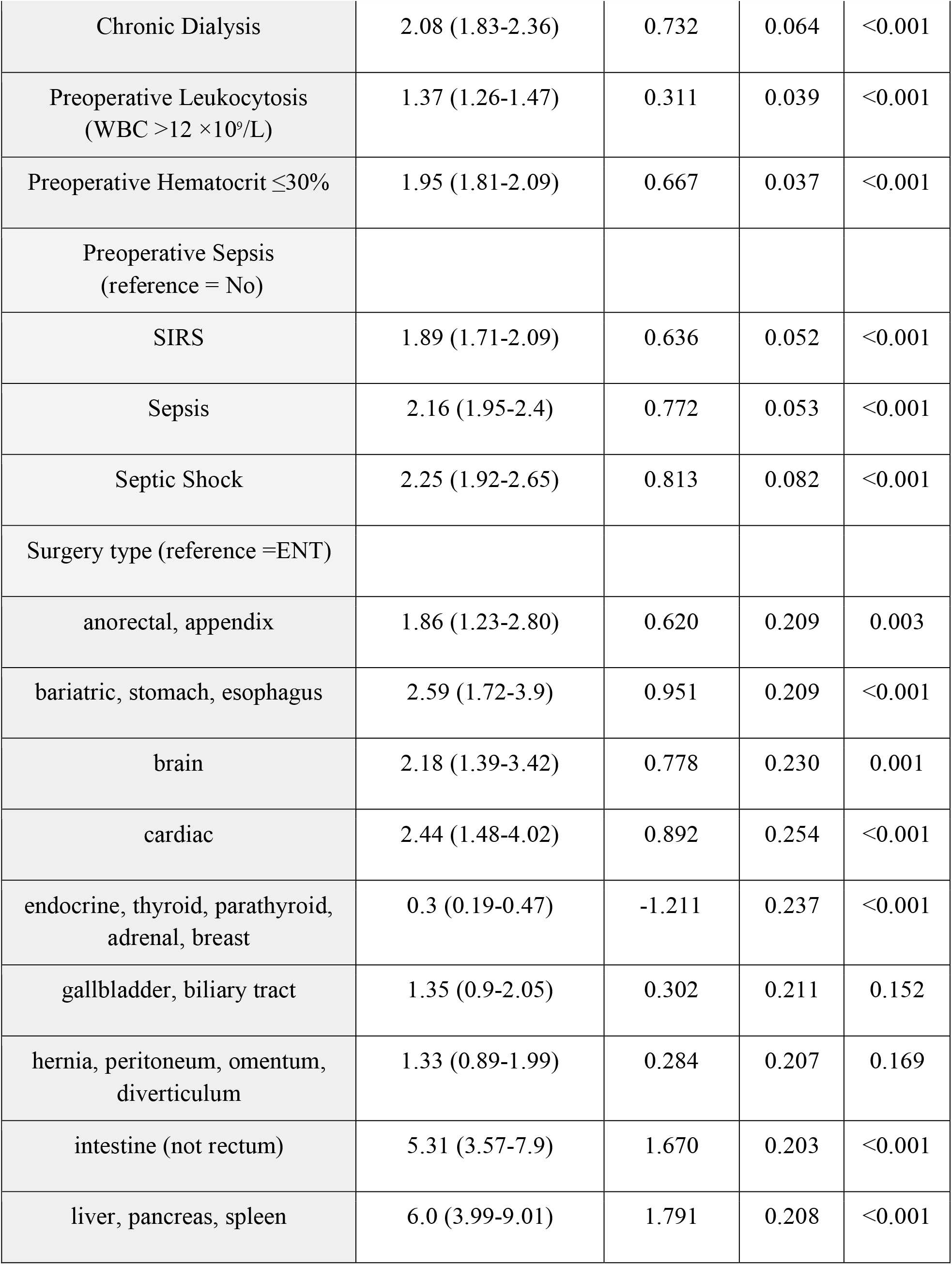
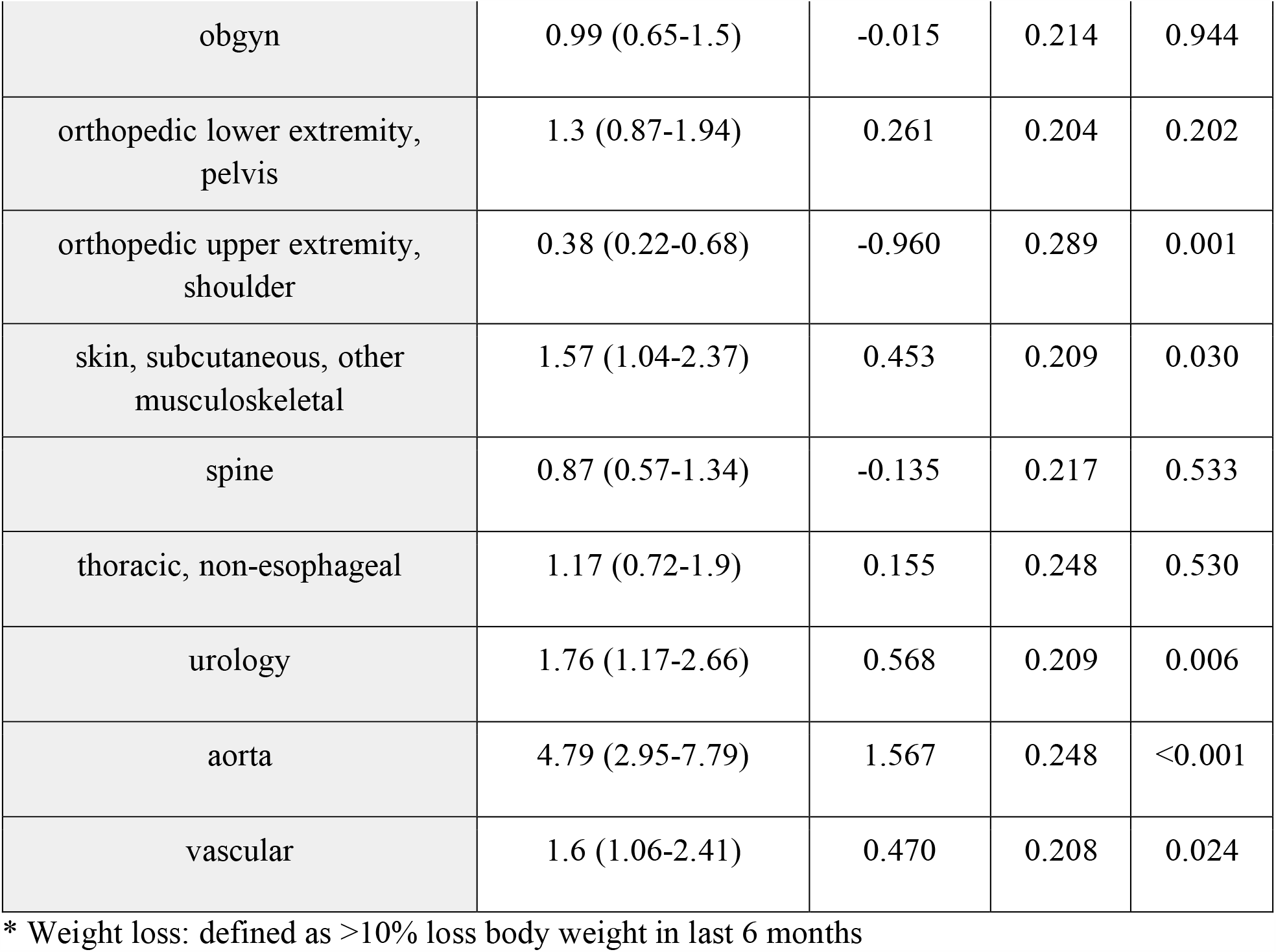
Adjusted odds ratio (OR) of *C. difficile* infection predictors

The risk model has excellent AUC (train cohort=0.804, test cohort=0.803).

### Inclusion of hospital length of stay as a predictor for postoperative *C. difficile* infection

**eTable I** in the **Supplement** shows 10 predictors for *C. difficile* infection including hospital length of stay. The hospital length of stay is significantly associated with *C. difficile* infection with P<0.001. The *C. difficile* risk model with 10 predictors including hospital length of stay had high predictive power (training cohort=0.841, testing cohort=0.838).

### Application of Postoperative *C. difficile infection* risk calculator

The probability of the occurrence of 30-day postoperative *C. difficile* infection is calculated using intercept and coefficients from table 2. The probability of case 3 was calculated using intercept and coefficients from Table I in the supplement. The model is available at http://cdiffrisk.herokuapp.com/. The web-based model was built using the Scikit-learn machine learning library (**eFigure 3 in the Supplement**). ^25^

<case 1>

60 year old male with a history of insulin dependent diabetes, ESRD (end stage renal disease) on dialysis, weight loss >10% in last 6 months, hematocrit 27%, leukocytosis (WBC 13×109/L), preoperative sepsis scheduled for hip surgery: 30-day postoperative *C. difficile* infection risk-5.79%.

<case 2>

60 year old female with a history of insulin dependent diabetes, no weight loss, no ESRD, no steroid, hematocrit 33%, no leukocytosis (WBC 8 ×109/L), no preoperative sepsis scheduled for hip surgery: 30 day postoperative *C. difficile* infection risk: 0.36%

As shown in these examples, the risk of postoperative *C. difficile* infection changes depending on comorbid conditions.

### Hospital day of CDI occurrences after surgery

The distribution of hospital days in which patients developed *C. difficile* infection is shown in **Figure 4**. Median hospital day of CDI occurrence was 9 days after surgery. 42.3% (N=4,091 out of 9,671) of CDI occurred post-discharge from 2015-2017 cohort. 73.5% of patients who developed CDI during the hospital admission were discharged to home, 16.8% went to a skilled care facility, and 6.5% to a rehabilitation facility.

**Figure 4.**
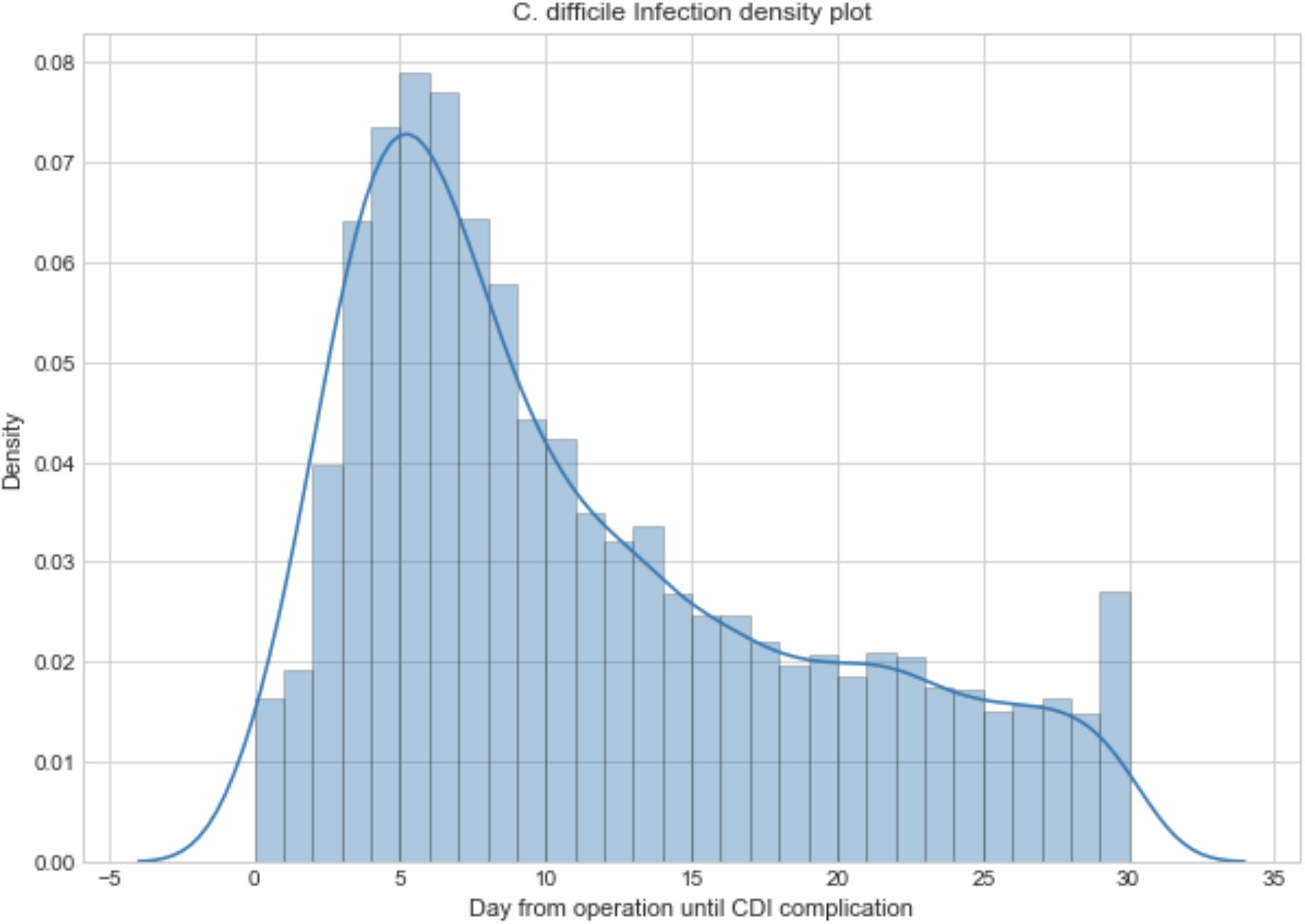
Distribution of hospital days from surgery until CDI complication.

## Discussion

Our study demonstrated that postoperative *C. difficile* infection was associated with high 30-day mortality and readmission rates similar to other studies. ^22^ Even though many hospitals have implemented measures to reduce *C. difficile* infection, the incidence of CDI remains high, thus novel approaches are needed to prevent this serious complication. In this large cohort study, we demonstrated a *C. difficile* prediction model which was developed and validated on over 2.4 million surgical patients from year 2015-2017. By accurately assessing postoperative risk of *C. difficile* infection, this risk assessment tool enables clinicians to apply different preventive measures according to the risk level. As was shown in this study, the risk of *C. difficile* infection increased with longer length of stay, therefore different preventive methods may need to be applied if patients’ hospitalization is longer than expected. Longer hospital length of stay likely increases the exposure to *C. difficile* spores. ^26,27^ Advanced age, weight loss, steroid use for chronic conditions and diabetes are related to impaired immune status, thus increasing the risk of CDI. ^26,28^

For patients with moderate to high risk of postoperative *C. difficile* infection, screening for *C. difficile* colonization and preventive isolation measures could be considered. Cleaning of patient rooms with ultraviolet disinfectant light may be useful. ^29^ Minimizing the use of proton pump inhibitors and unnecessary antibiotic exposure may help reduce the rate of CDI occurrence. ^30,31^ With a very high probability of postoperative *C. difficile* infection, the use of prophylactic probiotics and prophylactic antibiotics such as metronidazole or oral vancomycin could be a potential preventive measure, but these preventive strategies require further study to assess their efficacy and safety ^32^.

Our study is consistent with other studies which showed that preoperative infection and sepsis are associated with increased risk of *C. difficile* infection. ^22 14,33^

Previous studies showed a high incidence of CDI after appendectomy. ^34^ Our study showed that intestinal, vascular, liver, cardiac and stomach surgeries have higher rates of CDI than appendectomy. The disruption of normally protective colonic microflora layer with a procedure, nasogastric tube, enemas likely contributed to the high rate of CDI with intestinal and other gastrointestinal surgeries. ^26^

Our study also showed that a significant proportion (42.3%) of CDI occurred after hospital discharge. The most common discharge destinations of patients who developed CDI post-discharge were home (73.5%), followed by skilled care (16.8%) and rehabilitation facilities (6.5%).

Major strengths of our study: First, the model was built and validated across broad surgery types on over 2.4 million patients over three years. Second, the model was built from a cohort of national and international patients, so the model is more generalizable compared with models built from a small number of hospitals.

Limitations of our study: First, a history of prior *C. difficile* infection, recent exposure to antibiotics (including the use of perioperative antibiotics), and acid suppression therapy prior to surgery are important risk factors for *C. difficile* infection, ^9,15,35,36^ however our model did not include these factors because these were not parts of ACS-NSQIP database. ^31,37^ Second, variations of *C. difficile* infection testing among hospitals may impact testing results. ^38^ There is also an unknown correlation between hospital length of stay and CDI rate. Further analysis is needed to determine if a longer length of stay due to preoperative clinical factors led to a higher CDI rate or if this was secondary to longer exposure to pathogens and antibiotics.

## Conclusions

In conclusion, we developed and validated a clinically useful web-based predictive model for postoperative *C. difficile* infection. Our model enables clinicians to identify patients at high risk for *C. difficile* infection to implement individualized preventive measures. Patients with CDI were older, had more preoperative comorbidities such as disseminated cancer, COPD, ESRD, and insulin dependent diabetes. Independent risk factors included anemia, leukocytosis and SIRS, weight loss, intestinal surgery, aorta vascular surgery, liver-pancreas surgery, and longer length of stay. Further research is necessary to investigate the application of this risk model in clinical settings and revise current approaches to the prevention of postoperative *C. difficile* infection.

## Data Availability

The data is available at American College of Surgeon (ACS-NSQIP)

## Acknowledgments

Dr. Woo had full access to all the data in the study and takes responsibility for the integrity of the data and the accuracy of the data analysis.

## Funding/Support

*no funding*

*The American College of Surgeons National Surgical Quality Improvement Program and the hospitals participating in the ACS NSQIP are the source of the data used herein; they have not verified and are not responsible for the statistical validity of the data analysis or the conclusions derived by the authors*.

